# Estimation of mean pulmonary artery pressure by cardiovascular magnetic resonance four-dimensional flow and compressed sensing

**DOI:** 10.1101/2024.02.07.24302465

**Authors:** Goran Abdula, Pernilla Bergqvist, Jenny Castaings, Alexander Fyrdahl, Daniel Giese, Ning Jin, Frederik Testud, Peder Sörensson, Andreas Sigfridsson, Martin Ugander, David Marlevi

**Affiliations:** Dept. of Molecular Medicine and Surgery, Karolinska Institutet, Stockholm, Sweden; Dept. of Clinical Physiology, Karolinska University Hospital, Stockholm, Sweden; Magnetic Resonance, Siemens Healthcare, GmbH, Erlangen, Germany; Cardiovascular MR R&D, Siemens Medical Solutions USA, Inc, Cleveland, OH, USA; Siemens Healthcare AB, Malmö, Sweden; Dept. of Cardiology, Karolinska University Hospital, Stockholm, Sweden; Kolling Institute, Royal North Shore Hospital, St Leonards, Sydney, Australia; University of Sydney, Sydney, Australia; Institute for Medical Engineering and Science, Massachusetts Institute of Technology, Cambridge, MA, USA

**Author notes:** denotes corresponding author. denotes equal contribution as first author (Goran Abdula, Pernilla Bergqvist). denotes equal contribution as last author (Martin Ugander, David Marlevi).

## Abstract

**Background:** Four-dimensional (4D) phase-contrast cardiovascular magnetic resonance (CMR) allows for precise non-invasive estimation of mean pulmonary artery pressure (mPAP) by estimating the duration of pathological vortex persistence in the main pulmonary artery. This has previously been achieved with compressed sensing acceleration of a multiple two-dimensional (CS-M2D) flow sequence, but acquisition using a true time-resolved 3D excitation (CS-4D) offers theoretical advantages including spatiotemporal coherence. This study aimed to validate a state-of-the-art CS-4D sequence with a previously utilized CS-M2D sequence for estimating mPAP, and compare both to right heart catheterization (RHC).

**Methods:** The study included patients clinically referred for CMR (n=45), of which a subgroup (n=20) had prior mPAP of >16 mmHg confirmed by RHC. CMR was performed at 1.5T using CS-M2D and CS-4D sequences covering the main pulmonary artery. mPAP was estimated using a previously published linear relationship between vortex duration and mPAP. Agreement between CS-M2D and CS-4D estimates was quantified, including analysis of intra- and interobserver variabilities. The diagnostic performance of CS-M2D and CS-4D in predicting mPAP was further compared to gold-standard RHC.

**Results:** CS-M2D and CS-4D both had average scan durations under 3 minutes (175±36 and 135±34 seconds, respectively). Estimated mPAP by CS-4D and CS-M2D were strongly correlated (R^2^=0.93, p<0.001), with negligible mean±SD bias (0.0±2.7 mmHg) and good reproducibility. There was excellent agreement with RHC for both CS-M2D (R^2^=0.92, p<0.001, bias 0.6±3.1 mmHg) and CS-4D (R^2^=0.86, p<0.001, bias 1.1±4.5 mmHg).

**Conclusions:** CS-4D and CS-M2D sequences effectively yield interchangeable non-invasive estimations of mPAP, with excellent agreement compared to invasive RHC. They can both be acquired in a scan time applicable to clinical workflow, offering a promising tool for non-invasive mPAP estimation in clinical practice.

## Background

4D flow cardiovascular magnetic resonance (CMR) is a non-invasive imaging modality allowing for comprehensive assessment of full-field blood flow along arbitrary flow directions and throughout the entire cardiac cycle (1). The technique has been utilized across a variety of cardiovascular applications (2–5). In the setting of complex multidirectional flows, 4D imaging has shown advantages when compared to traditional 2D phase contrast imaging for both quantifying (6) and visualizing multidirectional flow (7). Moreover, 4D flow enables advanced hemodynamic assessment related to blood flow pattern, such as vorticity and helicity (8). In particular, recent studies have indicated the potential of using volumetric flow quantification using multiple stacked 2D (M2D) phase-contrast imaging with time-resolved three-directional velocity encoding to non-invasively quantify mean pulmonary artery pressure (mPAP) (9, 10) – a key diagnostic marker in diagnosing and prognosticating pulmonary hypertension (11).

Non-invasive estimation of mPAP depends on identification of vortical flow patterns in the main pulmonary artery, which can be performed in multiple ways. Previous work has utilized M2D imaging whereby 2D slices are stacked together to form a reconstructed 3D volume (12). By comparison, a true 4D acquisition, where the entire volume is acquired using a slab-selective excitation scheme and partition encoding, bears a number of theoretical advantages including temporal and spatial coherence, flexibility in acquiring isotropic voxels, and avoiding slice crosstalk. Furthermore, as non-invasive estimation of mPAP requires full coverage of the main pulmonary artery, its clinical adoption has been hindered by a relatively long scan time (13). The implementation of compressed sensing (CS) acceleration now promises acquisition in clinically acceptable scan times (14, 15). However, a direct comparison between M2D and true 4D flow have yet to be performed, and likewise, the accuracy of CS accelerated 4D (CS-4D) flow in detection of vortical flow and thereby estimation of mPAP remains unknown. Therefore, the aim of this study was to perform a comparison of CS accelerated M2D (CS-M2D) and CS-4D for estimating mPAP, with validation against right heart catheterization (RHC) in a sub-cohort of subjects with available invasive reference data.

## Materials and Methods

### Study Participants

The study cohort consisted of two groups. In the first group, thirty-five patients with referral for cardiac magnetic resonance (CMR) imaging who were suspected of having pulmonary hypertension (PH) were included. Reasons for suspected PH were known left ventricular systolic dysfunction or prior echocardiography revealing high systolic pulmonary artery pressure (sPAP). To validate CMR 4D flow estimated mPAP against the invasive reference standard RHC, a second group of 20 patients with mPAP >16 mmHg confirmed by RHC were enrolled. The cutoff of 16 mmHg was chosen since this cutoff represents the lowest pressure required for vortex formation (9). Patients with known contraindications for CMR, arrythmia, pacemaker, other cardiac implants, or valvular prosthesis were excluded. All subjects provided written informed consent, and the study was approved by the Swedish Ethical Review Authority (DNR: 2015/2106-31/1).

### CMR Imaging

CMR images were acquired using either a MAGNETOM Aera 1.5T (n=32) or MAGNETOM Sola 1.5T (n=23) (Siemens Healthineers AG, Erlangen, Germany). Anatomical and functional imaging was performed using breath-held cine imaging with balanced steady-state free precession (bSSFP) in both short and long axis views. Flow imaging of the main pulmonary artery (MPA) was acquired using compressed sensing (CS) accelerated time-resolved phase contrast imaging with three-directional velocity encoding, both using multiple 2D slices (CS-M2D) and volumetric excitation with partition encoding (CS-4D). The CS-M2D approach used 3 averages to reduce respiratory motion artifacts, whereas the CS-4D approach used either a crossed-pair or a pencil-beam respiratory navigator placed over the liver dome. **Table 1** shows a summary of imaging parameters used for both scanners.

**Table 1.** Sequence parameters of CS-M2D and CS-4D flow imaging at both scanner*s*.

| Parameters | CS-M2D |  | CS-4D |  |
| --- | --- | --- | --- | --- |
|  | Aera 1.5T | Sola 1.5T | Aera 1.5T | Sola 1.5T |
| Field of view (mm) | 340 | 340 | 320 | 320 |
| Voxel size (mm<sup>3</sup>) | 1.8 x 1.8 x 6.0 | 1.8 x 1.8 x 6.0 | 2.0 x 2.5 x 2.5 | 2.0 x 2.5 x 2.5 |
| Number of Slices | 10 | 10 | 30 | 30 |
| Velocity encoding (VENC, cm/s) | 90 | 90 | 90 | 90 |
| Bandwidth (Hz/pixel) | 449 | 449 | 558 | 558 |
| Flip angle (°) | 15 | 15 | 15 | 15 |
| Temporal resolution (ms) | 75.24 | 70.0 | 58.68 | 61.1 |
| Reconstructed time frames, n | 20 | 20 | 20 | 20 |
| Echo time (ms) | 4.10 | 3.61 | 2.55 | 3.0 |
| Repetition time (ms) | 6.27 | 5.83 | 4.89 | 5.09 |
| Slice coverage (mm) | 60 | 60 | 60 | 60 |
| Acceleration factor | 7.7 | 8.8 | 6.0 | 7.6 |
| Respiratory navigator type | - | - | Pencil beam | Cross-pair |

### Image Analysis

Left ventricular size, function, and mass were evaluated by manual segmentation of end-diastolic and end-systolic borders on the bSSFP cine 4-chamber view and short axis stacks using syngo.via (Siemens Healthineers AG, Erlangen, Germany). The basal dimension of the right ventricle (RV) at end-diastole (RVDd) and RV longitudinal function were determined by measuring tricuspid annular plane excursion (TAPSE) in the cine four-chamber view. Scan duration was recorded using a timer in a subgroup of patients, since actual scan duration was not stored in the dicom meta data.

For the hemodynamic image analysis, CS-M2D and CS-4D flow data were anonymized. The vortical blood flow in the MPA was assessed using a research software package (4D Flow, Siemens Healthineers AG, Erlangen,. Germany) in a randomized order. The datasets were pre-processed by applying background phase correction following cropping of spatially aliased structure if needed. The presence of pathological vortical blood flow was identified using multi-planar reconstructed 3-dimensional vector fields, see **Figure 1**. Vortex duration was defined as the percentage of the cardiac phases where vortical blood flow could be identified (12). Subsequently, mPAP was calculated using a previously determined empirical formula (9):

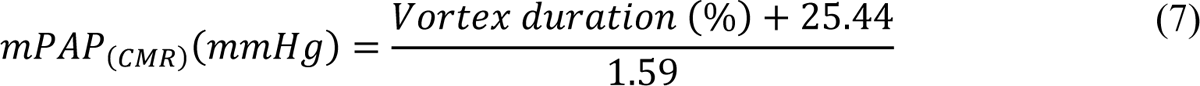

**Figure 1.**
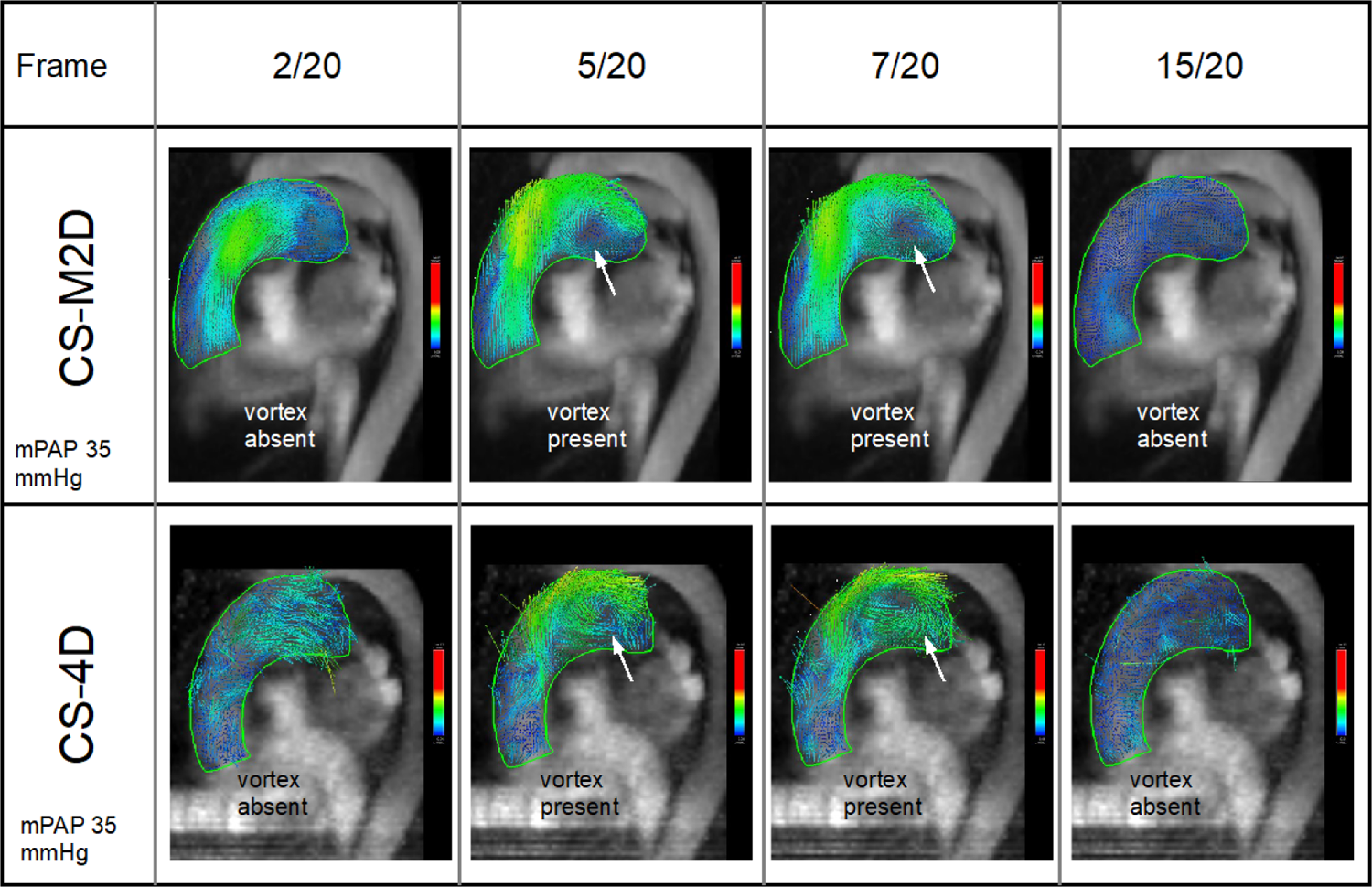
Vortical blood flow visualized in the main pulmonary artery in an oblique sagittal view for a patient with pulmonary hypertension (mPAP 35 mmHg by right heart catheterization), using CS-M2D (top row) and CS-4D (bottom row) across four representative time frames out of total 20 cardiac phases. The visualization is using multiplanar reformatted 3D velocity vector arrows color-coded for velocity. White arrow indicates a vortex.

The 4D flow analysis was performed for the first cohort by two independent readers (PB and a subset by GA), and for the second cohort by a single reader (GA). Analysis was consistently performed interchangeably on both CS-M2D and CS-4D datasets in a randomized fashion. Intra- and interobserver variability was assessed by selecting n=20 patients in the first cohort for which multiple readings were performed.

### Statistical analysis

Continuous variables were reported as mean ± standard deviation (SD) for normally distributed variables, while non-normally distributed variables were reported as median and interquartile range. Categorical variables were reported as frequencies and percentages. To quantify differences between CS-M2D and CS-4D measurements, two-tailed paired Student’s t-test was employed. Linear regression and Bland-Altman analysis were used to assess correlations and biases. Further, to quantify agreements between identified frames containing pathological MPA vortices, the Jaccard similarity coefficient was calculated as the mean of the index derived for absence, and presence of pathological MPA vortices, respectively.

Intra- and interobserver variability was evaluated by means of Bland-Altman statistics, calculating mean biases with standard deviations, along with the Jaccard similarity coefficient as per above. Statistical testing was performed using SPSS version 27 (IBM Corp., Armonk, NY, USA). A significance level of p<0.05 was utilized for all analyses.

## Results

Of the 55 prospectively included patients, 10 were excluded from analysis owing to the occurrence of arrhythmia during acquisition (n=4, affecting both CS-M2D and CS-4D), unrecoverable phase aliasing (n=2, affecting both CS-M2D and CS-4D), aborted ECG triggering (n=3, affecting CS-4D imaging), or post-scan reconstruction errors (n=1, affecting CS-4D). Excluded acquisition were equally distributed between the two utilized scanners (n=5 excluded from the Aera, n=5 excluded from the Sola). Demographics and CMR characteristics for the remaining 45 patients are summarized in **Table 2**.

**Table 2.** Patient demographics and clinical characteristics.

|  |  |
| --- | --- |
| Number of patients, n | 45 |
| Age, years | 55.6 ± 19.3 |
| Female, n (%) | 26 (58) |
| BMI, kg/m<sup>2</sup> | 24.8 ± 5.2 |
| BSA, m<sup>2</sup> | 1.9 ± 0.2 |
| Heart rate, beats per minute | 71 ± 13 |
| LV EDV, ml | 145 [112-191] |
| LV EDV index ml/m<sup>2</sup> | 80 [66-90] |
| LV ESV, ml | 59 [39-73] |
| LV ESV index, ml/m<sup>2</sup> | 31 [24-42] |
| LV SV, ml | 84 [68-103] |
| LV SV index ml/m<sup>2</sup> | 45 [38-53] |
| LV EF, % | 60 [54-64] |
| LV mass, g | 115 [87-144] |
| LV mass index, g/m<sup>2</sup> | 60 [50-70] |
| RVDd, mm | 46 [46-51] |
| TAPSE, mm | 24 [21-27] |

BMI = body mass index; BSA = body surface area; LV = left ventricular; EDV = end diastolic volume; ESV = end systolic volume; SV = stroke volume; RVDd = right ventricular dimension at end-diastole; TAPSE = tricuspid annular plane excursion.

### Diagnostic differentiation of pulmonary hypertension

Pulmonary hypertension (PH) defined as an estimated mPAP ≥20 mmHg was identified in 25/45 (56%) subjects by CS-M2D and 20/45 (44%) subjects by CS-4D. In 86% of the cases, both CS-M2D and CS-4D provided consistent classifications. Discrepancies arose in six subjects: five patients were classified with PH by CS-M2D, and one patient was classified with PH by CS-4D, see **Table 3**. For the subgroup who underwent reference standard invasive RHC (n=20), CS-M2D and RHC classifications agreed perfectly, confirming PH in 18 patients and ruling it out in two (mPAP <20 mmHg). Conversely, CS-4D exhibited discrepancies in three cases, two false positives and one false negative. An overview of the classification by CS-M2D vs CS-4D is given in **Figure 2**.

**Figure 2.**
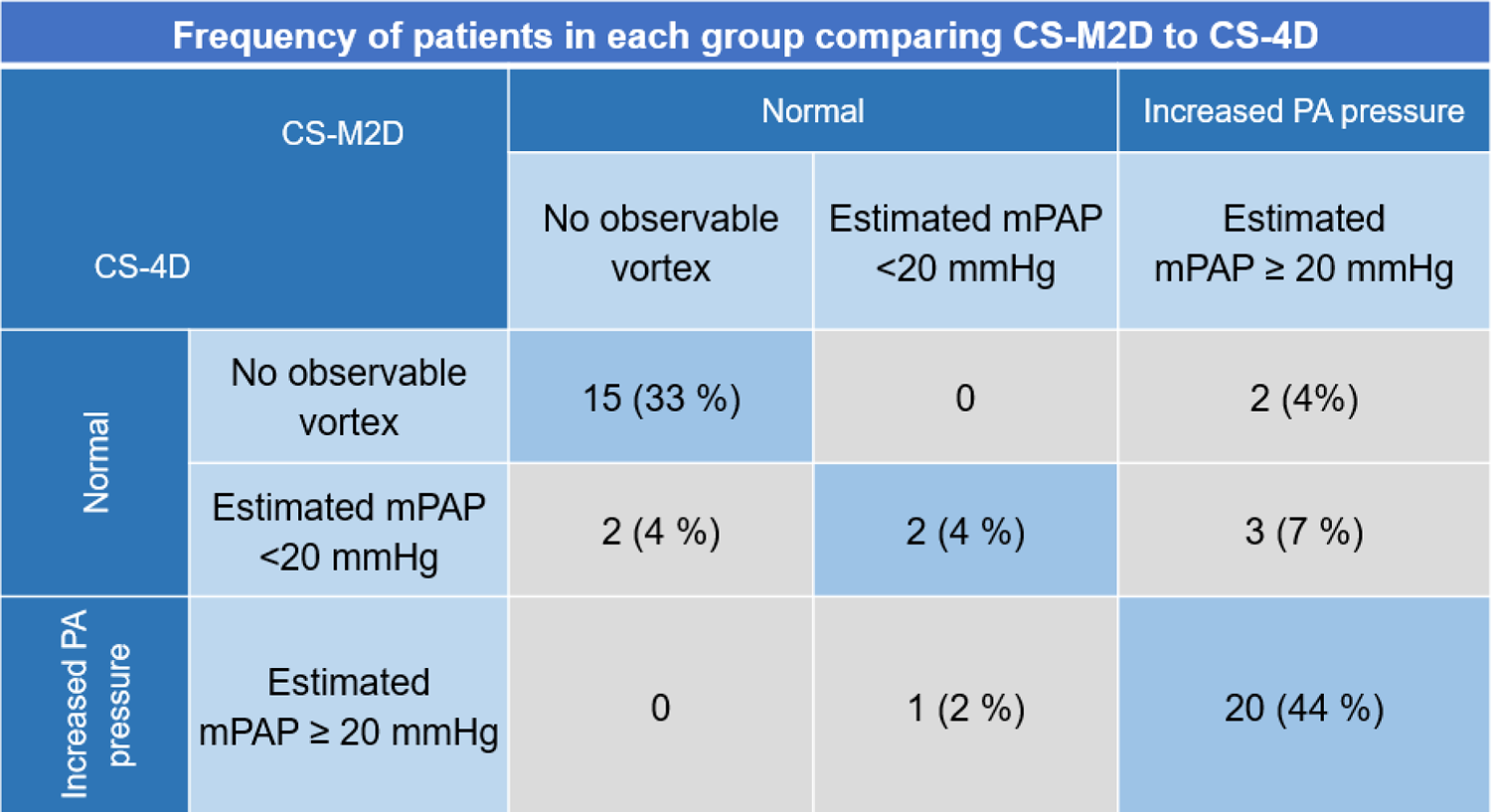
Distribution of pulmonary artery pressure diagnosis, comparing classification by PA vortex duration detected by CS-M2D and CS-4D.

**Table 3.** Identification of PH by CS-M2D vs CS-4D: mPAP > 20 mmHg.

| CS-4D | CS-M2D |  |  |
| --- | --- | --- | --- |
|  | Non-PH | PH | Total |
| Non-PH | 19 (42%) | 5 (11%) | 24 |
| PH | 1 (2%) | 20 (44%) | 21 |
| Total | 20 (44%) | 25 (56%) | 45 |

PH = pulmonary hypertension

### Quantification of mPAP

The time between RHC and CMR was 61 [40-142] days. Both CS-M2D and CS-4D approaches demonstrated excellent agreement for non-invasive estimation of mPAP with a high level of agreement (R²=0.93*, p<0.001*) and low bias (0.0±2.7 mmHg, **Figure 3**). There was no difference between estimated mPAP by CS-M2D and CS-4D in the overall cohort (24.3±9.9 vs. 23.7±9.2 mmHg, p = 0.15). The Jaccard similarity coefficient was 0.79 between mPAP estimated by CS-M2D and CS-4D across the entire cohort.

**Figure 3.**
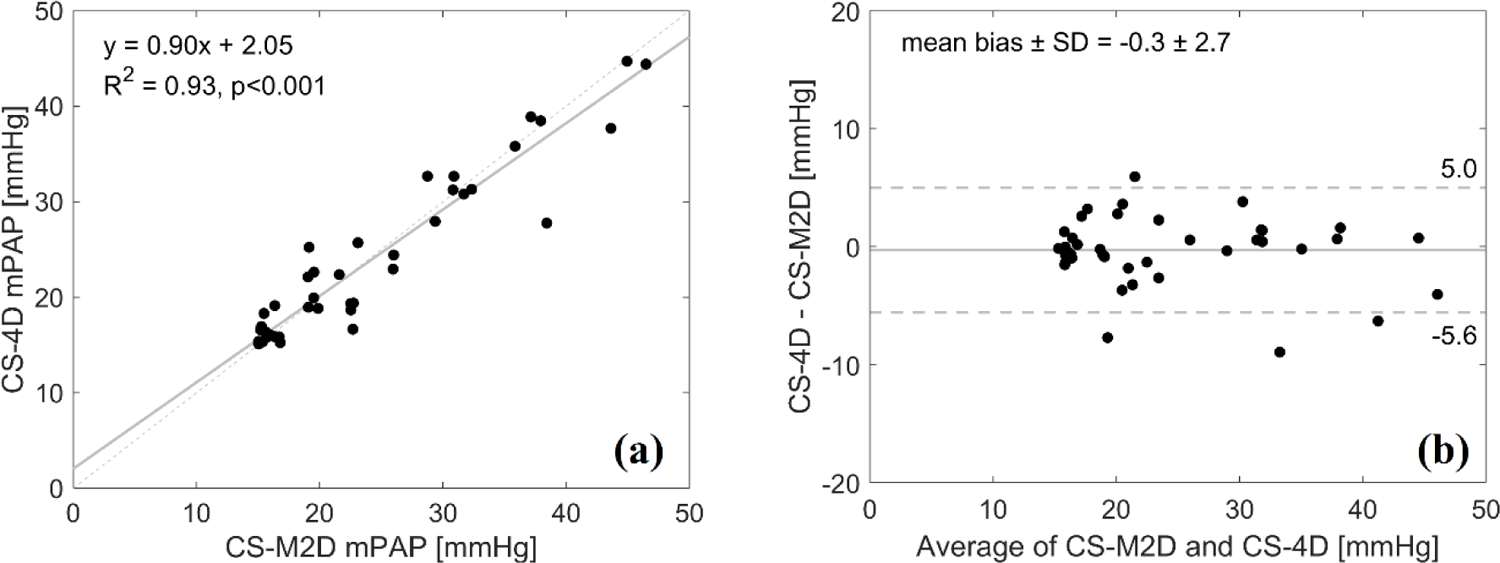
Linear regression (a) and (b) Bland-Altman Plots of the estimated mean pulmonary arterial pressures (mPAP) from vortex duration determined by CS-M2P and CS-4D.

### Comparison between estimated mPAP by CMR and RHC

Strong correlations were observed between mPAP measured by RHC and estimated by CS-M2D (R²=0.92*, p<0.001),* with an effective absence of any bias and good precision (0.6±3.1 mmHg, **Figure 4**). CS-4D also exhibited strong correlation with RHC (R²=0.86*, p<0.001)* and a similarly low mean bias and good precision (1.1±4.6 mmHg, **Figure 4**). Estimated mPAP did not differ between CS-M2D, CS-4D, and RHC (CS-M2D vs CS-4D: p=0.98; CS-M2D vs RHC: p=0.80; CS-4D vs RHC: p=0.59).

**Figure 4.**
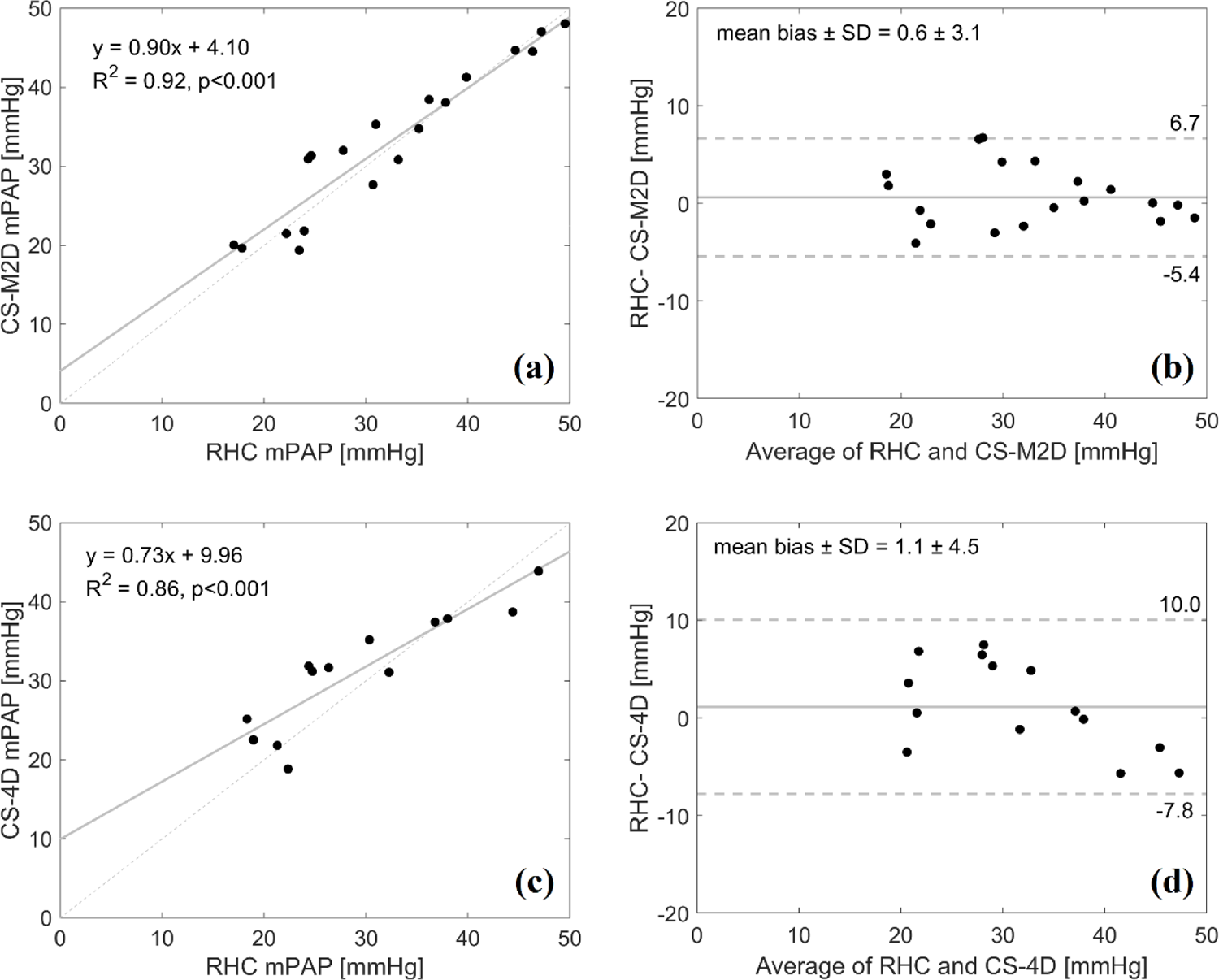
Linear regression and Bland-Altman Plots of mean pulmonary arterial pressures (mPAP) measured at right heart catheterization (RHC) and estimated mPAP by CS-M2D method (a-b) and estimated mPAP by CS-4D (c-d).

### Reproducibility analysis

From the intraobserver variability analysis, strong agreement was observed for both repeated CS-M2D (mean bias: 0.8±4.4 mmHg; Jaccard similarity index: 0.78) and CS-4D (mean bias: 0.8±2.0 mmHg; Jaccard similarity index: 0.79) readings, respectively. For the interobserver variability analysis, similar negligible bias was observed although with higher variation for both repeat CS-M2D (mean bias: 0.3±9.0 mmHg; Jaccard similarity index: 0.54) and CS-4D (mean bias: 1.6±7.7 mmHg; Jaccard similarity index: 0.55) readings, respectively. An overview of the reproducibility readings is provided in **Figure 5**.

**Figure 5.**
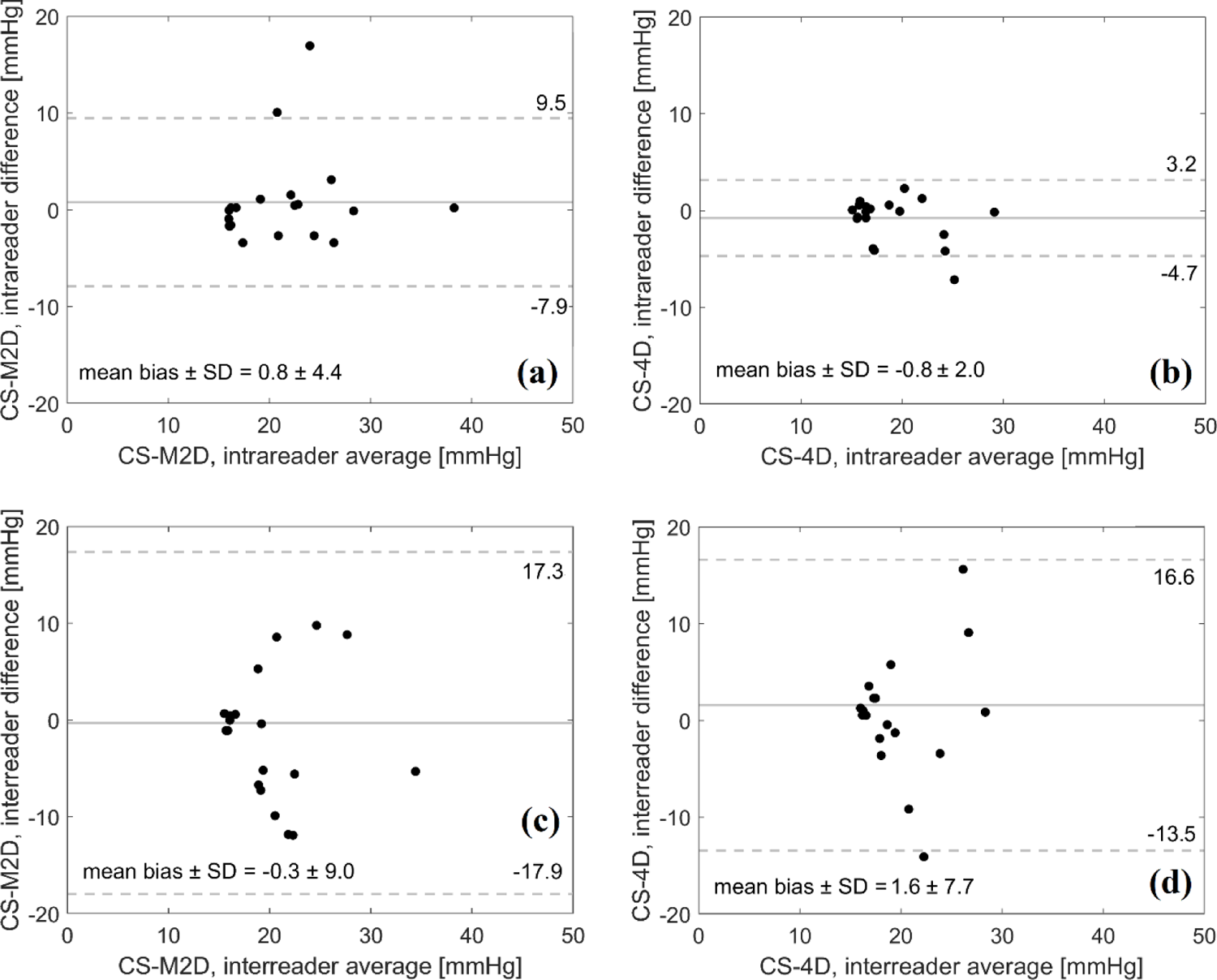
Bland-Altman Plots of mean pulmonary arterial pressures (mPAP) estimated by CS-M2D and by CS-4D, showing intra-(a-b) and interobserver (c-d) variability across a randomly selected subset of n=20 patients.

### Scan time

With estimates taken from a subgroup of 8 patients, average scan duration for CS-M2D and CS-4D were 175±36 s and 135±34 s, respectively.

## Discussion

The main finding of the current study is that both CS-M2D and CS-4D imaging approaches using CMR yield interchangeable results that both represent accurate estimations of mPAP when validated against invasive RHC. These results further highlight the potential of CMR as a non-invasive alternative for diagnosis of pulmonary hypertension; in-line with previous findings in literature (9, 16, 17).

In previous studies, CMR-based estimation of mPAP has been performed using the M2D approach without CS acceleration (9, 10, 12). Whilst a recent study showed that CS acceleration had non-inferior performance (16), the current study represents the first head-to-head comparison between CS-M2D and CS-4D, and the first comparison of these techniques to invasive RHC. Here, we aimed to replicate the previously published CS-M2D sequence, with the CS-4D representing a state-of-the-art comparator. As highlighted in **Table 1**, this resulted in a number of differences relating to effective voxel size, temporal resolution, and overall acquisition time. Specifically, CS-M2D used a voxel size of 1.8×1.8×6.0 mm^3^ as compared to CS-4D at 2.0×2.5×2.5 mm^3^. As such, flow coverage by CS-4D is almost twice as dense as compared to CS-M2D in the slice direction, with only minor differences in in-plane resolution. CS-4D would thus have the theoretical ability to quantify finer flow details compared to CS-M2D. Furthermore, there are slight differences in temporal resolution in the utilized acquisition settings, and the nature of CS-M2D imaging involves spatially contiguous but temporally disparate slice acquisitions. This presents theoretical challenges for the CS-M2D approach, particularly in transient or complex anatomies and flows. However, the current study highlights the effectively equivalent capabilities of CS-4D and CS-M2D for non-invasive estimation of mPAP despite these theoretical differences and challenges.

Early detection of PH is associated with improved prognosis (18), however, the majority of PH patient diagnosed present at an advance stage (NYHA class III and IV), leaving early detection a remaining and urgent clinical challenge (19). Early detection is typically attempted using echocardiography. However, it has been shown that identifying PH by 4D flow analysis by CMR has twice the diagnostic yield compared to echocardiography (20), and this increased diagnostic performance has been confirmed compared to RHC (17).

Furthermore, identifying PH is a central component of assessing diastolic dysfunction, which plays a central role in the diagnosis and therapeutic evaluation in heart failure with preserved ejection fraction (HFpEF). Indeed, estimation of mPAP by 4D flow analysis can be used to perform grading of diastolic dysfunction by CMR, and this has shown excellent agreement compared to echocardiography (21). These described clinical applications of 4D flow analysis by CMR to date have all used M2D acquisition without CS acceleration, and the results of the current study now provide the field with important confidence to use CS to drastically reduce acquisition time from an average of 9 minutes to under 3 minutes, which provides valuable improvements in clinical throughput capacity for CMR imaging facilities.

Insights into clinical utility were also provided by the reproducibility study. As presented in **Figure 5**, negligible bias was reported in an intraobserver setting, and in the interobserver setting observer bias was kept below 1.6 mmHg across all subjects and acquisition sequences, respectively. Nevertheless, increased variance was observed in the interobserver setting; a finding aligning with the nature of multiple observers, but still highlighting challenges associated with manual image interpretation. Recent developments on semi-automatic or even fully automatic vortex detection in the setting of pulmonary flow imaging has shown high clinical potential (22, 23), herein offering a promising path towards mitigating reader bias and maintaining accurate estimation of performance.

The current study has some limitations that need to be acknowledged. First, a relatively small number of patients were included, in particular in the subgroup that underwent RHC. As such, further validation in larger cohorts could be of benefit, and would aid in the understanding of how the current results can generalize into other centers or across larger and differently composed populations. That said, it has already been shown that estimating mPAP with the M2D approach is excellently accurate and robust across all subtypes of PH (9).

Second, pathological vortex detection as performed in the current study is currently a manually performed method, which is not only time consuming, but also leaves room for observer variations as highlighted in the reproducibility study. As noted above, efforts to reduce observer dependence have recently been presented through incorporation of semi-automatic or fully automatic vortex detection algorithms (22, 23). A such, the current results provide the basis and confidence for understanding accuracy and sequence performance, from which further optimization can be envisioned.

## Conclusion

CS-accelerated CMR 4D flow analysis provides means for accurate and clinically feasible non-invasive assessment of mPAP using either CS-M2D or CS-4D approaches, opening for a more accessible way of diagnosing PH compared to invasive catheterization.

## List of abbreviations

4D: Four-dimensional

CMR: cardiovascular magnetic resonance

mPAP: mean pulmonary artery pressure

CS: compressed sensing

CS-M2D: compressed sensing multiple two-dimensional (flow sequence)

CS-4D: compressed sensing time-resolved three-dimensional (flow sequence)

RHC: right heart catheterization

bSSFP: balanced steady-state free precession

MPA: main pulmonary artery

RV: right ventricle

TAPSE: tricuspid annular plane excursion

## Declarations

### Ethics approval and consent to participate

All subjects provided written informed consent, and the study was approved by the Swedish Ethical Review Authority (DNR: 2015/2106-31/1).

### Consent for publication

Not applicable.

### Availability of data and materials

The datasets used and/or analysed during the current study are available from the corresponding author on reasonable request.

### Author’s contributions

D.M. and M.U. conceived of the study, with G.A., P.B., M.U. and D.M. involved in study design, patient recruitment, and primary image and statistical analysis. D.G., N.J., and F.T. were involved in sequence acquisition design, with A.F., A.S., and J.C. leading local implementation and guidance on image acquisition and analysis. D.M., M.U., A.S., and P.S. supervised P.B and G.A., taking part in analysis and preliminary data assessment. All authors were involved in manuscript drafting, and all authors approved of the final manuscript.

## Acknowledgement

Not applicable.

